# Changing Pattern of Paediatric Endocrinology Referrals over two decades: A Retrospective Study from a Tertiary Centre in Western India

**DOI:** 10.1101/2025.01.03.25319937

**Authors:** Khadilkar Vaman, Karishma K. Bhade, Sonali Wagle-Patki, Anuradha V. Khadilkar

## Abstract

**Introduction:** Understanding the paediatric endocrinology referral pattern is important for primary care clinicians and paediatric endocrinologists to optimise patient care, facilitate continuous medical education and to upgrade resources. This study analysed the pattern of these referrals over a year, the change in referral trends 2 decades apart and the discrepancy between the referral reasons and final diagnoses.

**Methods:** A retrospective analysis was conducted on demographic details and referral reasons to paediatric endocrine clinic in a tertiary care hospital in Western India at 2-time intervals 2 decades apart (2002-03 and 2022-23). The referral reasons were categorised into 14 classes as per International classification of paediatric endocrinology diagnoses (ICPED) 2016.

**Results:** Data of 2595 patients (920 from 2002-03 and 1675 from 2022-23) was studied. The commonest reason for referral was Short Stature with no gender bias. Disorders of Puberty and Obesity were the 2nd and 3rd commonest reasons for referral. There was almost a 2-fold rise in the total number of referrals over 2 decades with a significant rise in females referred for Short Stature and Disorders of Puberty. There was a discrepancy between the final diagnosis and referral reason, predominantly in patients referred for Micropenis, Gynaecomastia and Obesity

**Conclusion:** We report paediatric endocrine referral pattern over two decades, revealing a shift in the number of referrals but not a major shift in the referral reasons. A gap exists in recognizing symptoms and possible cause at the primary care level. These findings highlight the need for focused medical education and awareness among primary care clinicians.

## Introduction

Disorders of hormonal imbalance and stature are common causes of concern for parents and children. ^[1–3]^ As demographic changes also change the disease frequency and patterns; it is necessary to study and document changing disease spectrum over time that gets referred to any speciality unit. There is scarcity of data on the referral patterns of paediatric endocrine disorders in Indian literature. Over the past few decades, incidence of precocious puberty and type 1 diabetes is reported with increasing frequency. Further, as India goes through the phase of economic and nutritional transition, disorders such as obesity are also on the rise.

^[4]^ The initial contact for a child’s health issues typically begins with a general paediatrician. A general paediatrician thus plays a critical role in primary care of a child, including identifying and recommending specialty consultations as needed. Effectively addressing these concerns and ensuring appropriate referrals to a paediatric endocrinologist when necessary are essential for delivering timely and effective treatment.^[5]^ These understanding aids in efficiently organising and prioritising outpatient clinics, facilitating continuous medical education for primary care providers, and preventing the loss of referrals over time.

Among all the endocrine disorders, referral pattern and aetiology of short children has been well documented in Indian studies, however, there are very few Indian studies (or studies from other countries) describing the pattern of overall paediatric endocrine referral.^[6,7]^ In 2020, Bellotto et al conducted a retrospective study analysing referral patterns in paediatric endocrinology over a six-year period at a single tertiary centre in Italy. ^[8]^ As the referral frequency and pattern seems to have changed over a period of a few decades, the knowledge of patterns and features of referrals is crucial to guide primary care providers as well as paediatric endocrinologists.

The purpose of this study was thus to retrospectively analyse changing patterns of paediatric endocrine referrals made to a tertiary level centre in Western India two decades apart (2002-2003 vs 2022- 2023) and to compare the reasons for referral with the final diagnosis by a paediatric endocrinologist. Our specific objectives were:

1) To study the paediatric endocrine referral pattern over a year in a tertiary care hospital in Western India (2022-23)
2) To assess the change in referral trends 2 decades apart (2002-2003 vs 2022- 2023)
3) To study the discrepancy between reasons for referral and the final diagnosis made by the paediatric endocrinologist

## Material and Methods

We retrospectively analysed data on patients referred to the paediatric endocrine outpatient unit of our tertiary care hospital in Western India at two-time intervals 20 years apart. First dataset is from January 2002 to December 2003 and the second dataset is from January 2022 to December 2023.

The deidentified data were procured from the treating consultant’s electronic records. These patients were referred by a paediatrician or a family physician. Information regarding the reason for referral, sex and date of birth was retrieved at both time intervals. Patients were categorised based on the referral reasons into 14 classes as per International classification of paediatric endocrinology diagnoses (ICPED) 2016.^[9]^ These classes are Short Stature, Tall Stature, Puberty, Sex Development and Gender, Obesity, Pituitary Gland, Hypothalamus and Central Nervous System, Thyroid Gland, Adrenal glands, Testes and male reproductive tract, Ovaries and female reproductive tract, Glucose and lipid metabolism, Calcium and phosphate metabolism, Salt and water regulation, Syndrome with endocrine features. These main classes were further sub-categorised as per ICPED 2016. The final diagnosis was made based on the clinical history and examination, anthropometric parameters and investigations. The electronic records were exported to an excel sheet. Analysis was performed with IBM SPSS version 29 (SPSS, Chicago, IL, USA). Data are presented as frequencies and percentages. The differences in the proportions of referral patterns over two decades among males and females were calculated using the z-test for proportion. The Institutional Ethics Committee granted a waiver to analyse data that had been appropriately deidentified to ensure privacy and confidentiality.

## Results

Data on a total of 2595 patients (1675 from 2022-23 and 920 from 2002-03) were analysed. Out of 1675 patients in 2022-23, 770 were males (median age-11.1 years, IQR-7.5-13.7) and 905 were females (median age-10.4, IQR-7.2-13.5). The most common five referral categories were Short Stature (59.6%), Puberty (13%), Obesity (7.5%), Thyroid gland (5.6%) and Testes & Male Reproductive Tract (3.8%) (Figure 1a).

**Figure I.**
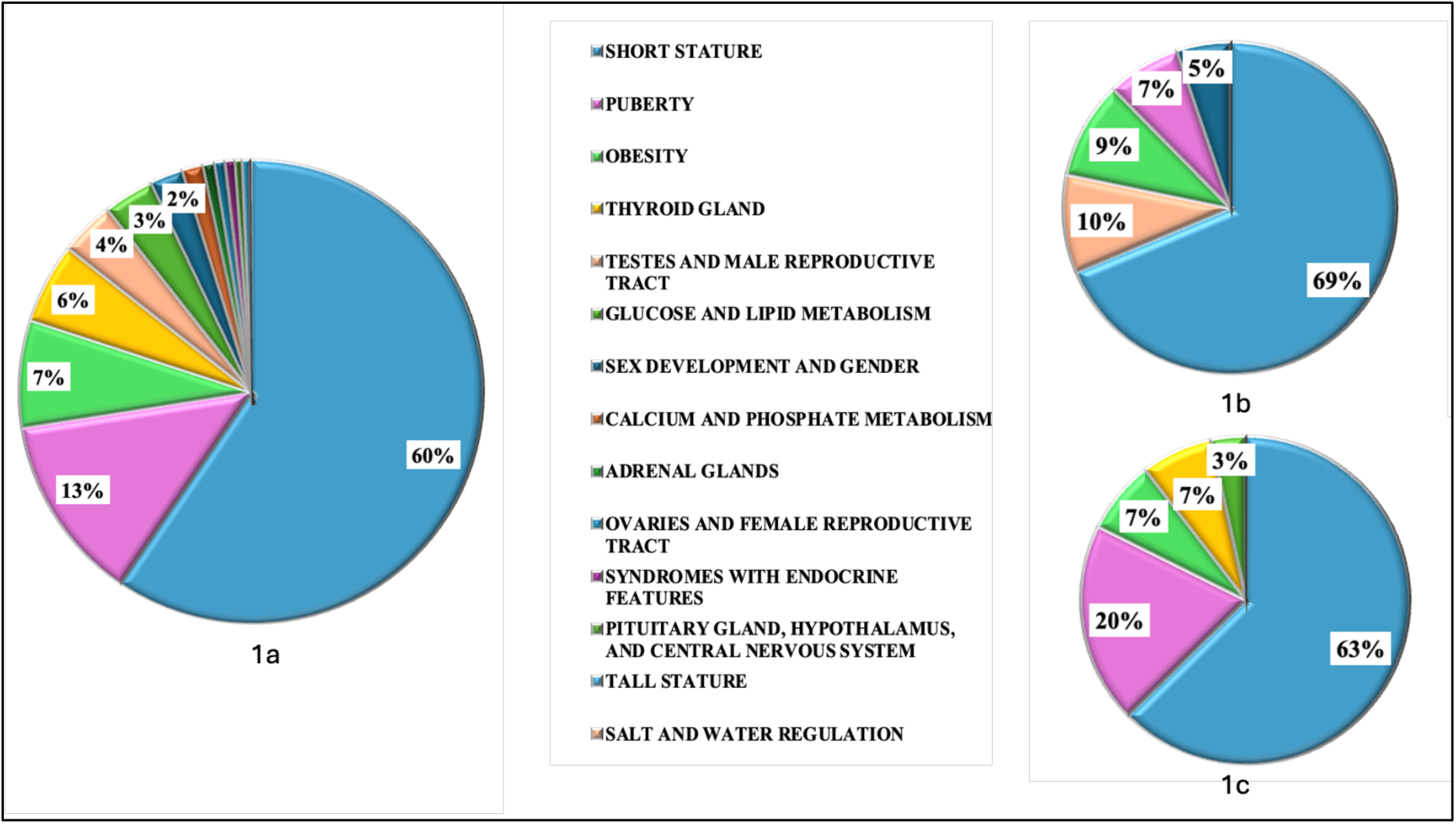
Distribution of referral categories as per proportions in total patients(1a), males (1b) and females (1c). The most common 5 referral categories are Short Stature, Disorders of Puberty, Obesity, Disorders of Thyroid Gland and Disorders of Testes and Male Reproductive Tract. Short Stature is the most common reason for referral in both males and females followed by Disorders of Testes and Male Reproductive Tract in males and Disorders of Puberty in females.

Sex wise distribution of the most common 5 main categories is as shown in figure 1b and 1c. Short Stature was the commonest reason for referral in both males (59.7%) and females (59.6%). Disorders of Testes and Male Reproductive Tract (8.3%) and disorders of Puberty (18.7%) were the second most common categories in males and females respectively. This was followed by Obesity (8.2%), Puberty (6.2%) and disorders of Sex Development and Gender (4.5%) in males and Obesity (6.9%), disorders of Thyroid Gland (6.7%) and disorders of Glucose and Lipid Metabolism (3.2%) in females.

### Subcategories of the most common five classes (Figure 2)

#### Short Stature (Fig 2a)

Among 999 patients with Short Stature, 49.6% were diagnosed to have Idiopathic short stature (Familial Idiopathic short stature (FSS)-22.5%, Non-familial idiopathic short stature-14.5%, Constitutional delay in growth and puberty (CDGP)-11.3%, Delayed puberty-1.3%), 29.4 % had Primary growth failure (Small for gestational age with failure to catch up growth-17.8%, Syndromic short stature-6.1%, Skeletal dysplasia-5.5%,), 19% had Secondary growth failure (Endocrine causes-8.3%, Nutritional causes-6.5%, Chronic illness-3%, Other skeletal defects-1.1%) and 1.1 % had Normal stature.

**Figure II.**
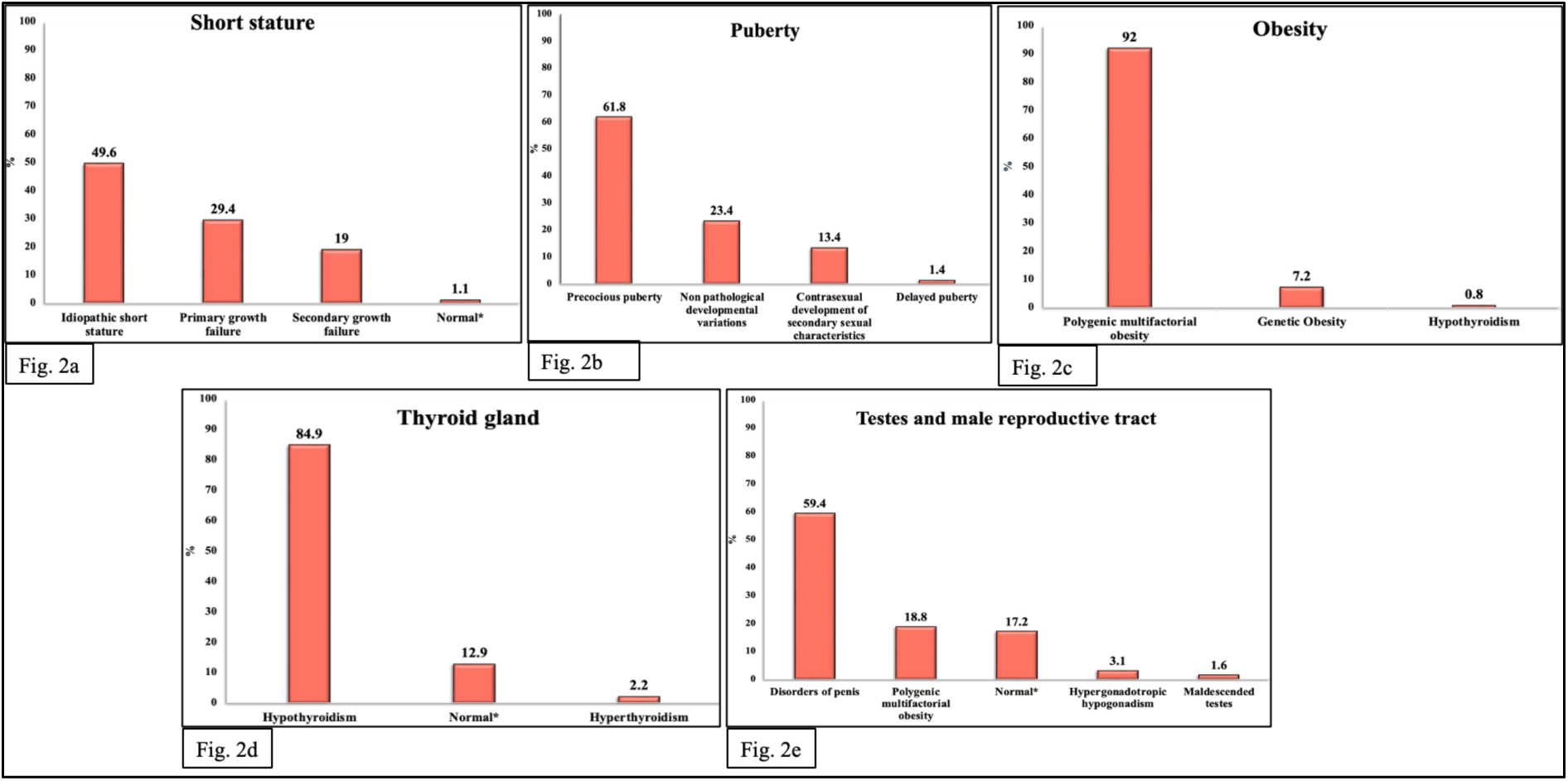
Frequency distribution of subcategories in each of the most common 5 referral categories. *Denotes the subjects who were referred for a particular disorder but found to have no pathological condition and were normal Fig 2a: The most common cause of Short Stature was Idiopathic short stature (ISS) [FSS-22.5%, Non-familial ISS-14.5%, CDGP-11.3%, Delayed puberty-1.3%] Fig 2b: The most common cause of Disorders of Puberty was Precocious puberty Fig 2c: The most common cause of Obesity was Polygenic multifactorial obesity Fig 2d: The most common cause of Disorders of Thyroid Gland was Hypothyroidism. The findings of 12.9% patients referred for suspected disorders were normal Fig 2e: The most common cause of Disorders of Testes and Male Reproductive tract was Disorders of penis. 17.2% patients referred with suspected pathology were normal

#### Puberty (Fig 2b)

Among 217 patients referred for disorders of Puberty, 61.8% had Precocious puberty, 23.4% had Non-pathological developmental variations, 13.4% had Contra-sexual development of secondary sexual characteristics in which all the patients were males with puberty gynaecomastia and 1.4% had Delayed puberty.

#### Obesity (Fig 2c)

Out of 125 patients referred for obesity, 92% had Polygenic, multifactorial obesity, 7.2% Genetic obesity and 0.8% had Hypothyroidism.

#### Thyroid gland (Fig 2d)

Out of 93 patients with disorders of Thyroid gland, 84.9% had Hypothyroidism, 12.9% had no clinical or biochemical pathology and hence labelled Normal, and 2.2 % had Hyperthyroidism.

#### Testes and Male Reproductive Tract (Fig 2e)

Sixty-three males were referred for disorders of Testes and Male Reproductive Tract. Out of these, 78.2% had disorders of Penis, 17.4% had Normal variants, 3.1% had Hypergonadotropic hypogonadism and 1.6% had Maldescended testis.

### Change in the pattern of referrals over 2 decades (Fig. 3) (Table 1)

The total number of referrals and outpatient load had increased from 2002-03 to 2022-23 by 82%. In males, the number increased by 54% and in females it increased by 115%. The referrals for Obesity were significantly decreased in males. However, the referrals for Short Stature and Puberty were increased but decreased for Obesity, disorders of Ovaries and Female Reproductive Tract and Thyroid Gland.

**Figure III.**
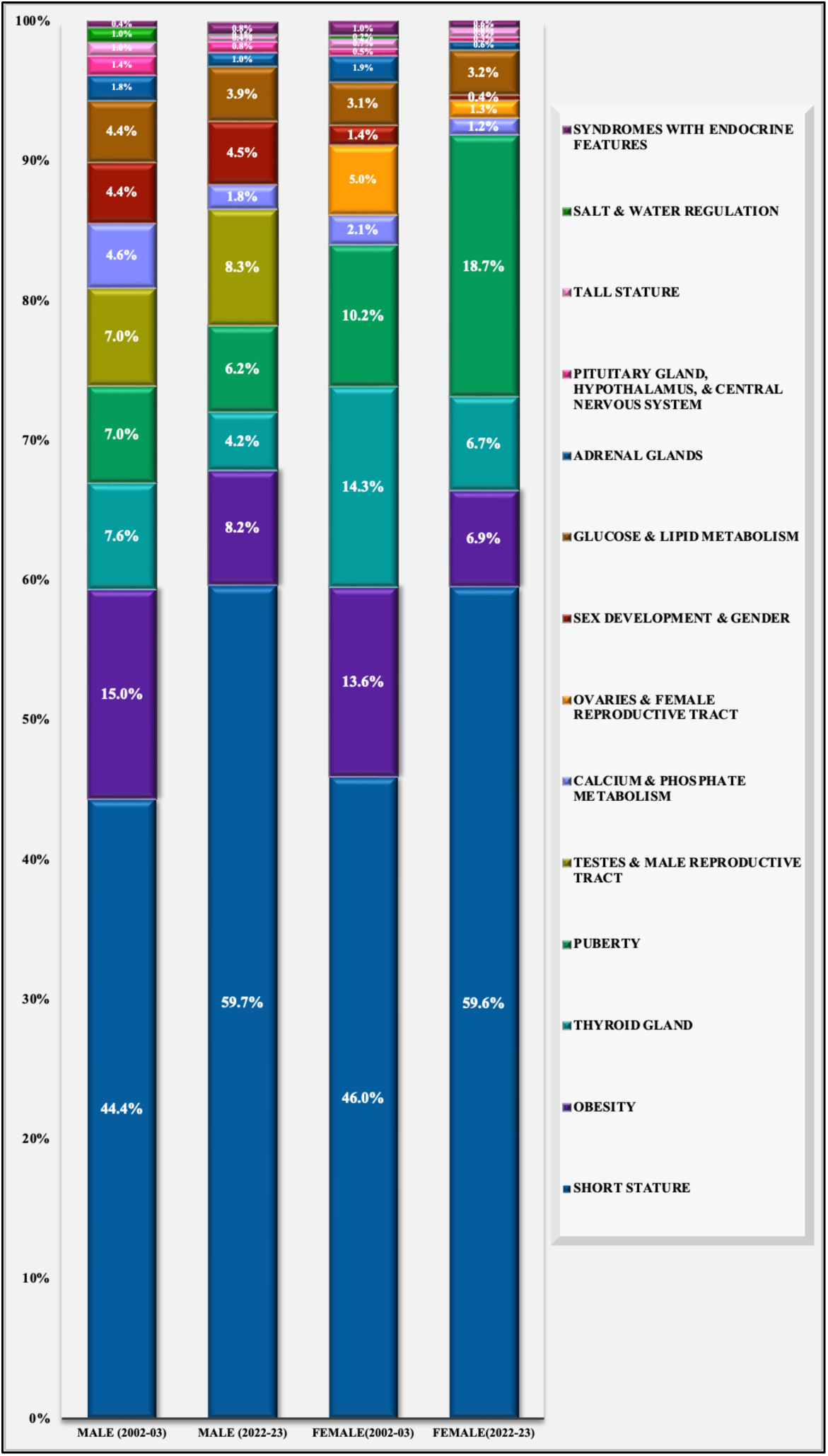
Comparison of Referral categories in males and females in 2002-03 and 2022-23 Distribution of referral reasons among males and females as per proportions 2 decades apart

**Table I.** Change in the pattern of referral categories over 2 decades in total patients, males and females. Values are in n (%), comparison between two groups was done using z test for proportion

|  | Total |  |  | Males |  |  | Females |  |  |
| --- | --- | --- | --- | --- | --- | --- | --- | --- | --- |
|  | 2002-03 | 2022-23 | *P* value | 2002-03 | 2022-23 | *P* value | 2002-03 | 2022-23 | *P* value |
| Referral Categories | n=920 | n=1675 |  | n=500 | n=770 |  | n=420 | n=905 |  |
| Short Stature | 415<br>(45.1 %) | 999<br>(59.6 %) | <0.001 | 222<br>(44.4 %) | 460<br>(59.7 %) | 0.003 | 193<br>(46 %) | 539<br>(59.6%) | <0.001 |
| Tall Stature | 8<br>(0.9 %) | 8<br>(0.5 %) | 0.224 | 5<br>(1 %) | 3<br>(0.4 %) | 0.180 | 3<br>(0.7 %) | 5<br>(0.6 %) | 0.723 |
| Puberty | 78<br>(8.5 %) | 217<br>(13 %) | 0.001 | 35<br>(7 %) | 48<br>(6.2 %) | 0.584 | 43<br>(10.2 %) | 169<br>(18.7%) | <0.001 |
| Sex Development and Gender | 28<br>(3 %) | 39<br>(2.3 %) | 0.272 | 22<br>(4.4 %) | 35<br>(4.5 %) | 0.902 | 6<br>(1.4 %) | 4<br>(0.4 %) | 0.053 |
| Obesity | 132<br>(14.3%) | 125<br>(7.5 %) | <0.001 | 75<br>(15 %) | 63<br>(8.2 %) | <0.001 | 57<br>(13.6 %) | 62<br>(6.9 %) | <0.001 |
| Pituitary Gland, Hypothalamus, And Central Nervous System | 9<br>(1 %) | 9<br>(0.5 %) | 0.195 | 7<br>(1.4 %) | 6<br>(0.8 %) | 0.283 | 2<br>(0.5 %) | 3<br>(0.3%) | 0.689 |
| Thyroid Gland | 98<br>(10.7%) | 93<br>(5.6 %) | <0.001 | 38<br>(7.6 %) | 32<br>(4.2 %) | 0.008 | 60<br>(14.3 %) | 61<br>(6.7 %) | <0.001 |
| Adrenal Glands | 17<br>(1.8 %) | 13<br>(0.8 %) | 0.014 | 9<br>(1.8 %) | 8<br>(1 %) | 0.249 | 8<br>(1.9 %) | 5<br>(0.6 %) | 0.020 |
| Testes And Male Reproductive Tract | 35<br>(3.8 %) | 64<br>(3.8 %) | 0.983 | 35<br>(7 %) | 64<br>(8.3 %) | 0.394 | - | - | - |
| Ovaries And Female Reproductive Tract | 21<br>(2.3 %) | 12<br>(0.7 %) | 0.001 | - | - | - | 21<br>(5 %) | 12<br>(1.3 %) | <0.001 |
| Glucose And Lipid Metabolism | 35<br>(3.8 %) | 59<br>(3.5 %) | 0.710 | 22<br>(4.4 %) | 30<br>(3.9 %) | 0.658 | 13<br>(3.1 %) | 29<br>(3.2 %) | 0.916 |
| Calcium And Phosphate Metabolism | 32<br>(3.5 %) | 25<br>(1.5 %) | 0.001 | 23<br>(4.6 %) | 14<br>(1.8 %) | 0.004 | 9<br>(2.1 %) | 11<br>(1.2 %) | 0.197 |
| Salt And Water Regulation | 6<br>(0.7 %) | 1<br>(0.1 %) | 0.005 | 5<br>(1 %) | 1<br>(0.1 %) | 0.027 | 1<br>(0.2 %) | 0<br>(0 %) | 0.142 |
| Syndromes With Endocrine Features | 6<br>(0.7 %) | 11<br>(0.7 %) | 0.991 | 2<br>(0.4 %) | 6<br>(0.8 %) | 0.404 | 4<br>(1 %) | 5<br>(0.6 %) | 0.409 |

Short Stature was the most frequent reason for referral overall and across both sexes during the two time periods. Notably, the proportion of referrals for Short Stature increased significantly, from 45.1% in 2002–2003 to 59.6% in 2022– 2023. While this upward trend was observed in both males and females, statistical significance was evident only in females.

The total number of patients referred for disorders of Puberty had increased from 8.5% to 13% (*P*<0.001). This increase was predominantly in females from 10.2% to 18.7%(*P*<0.001). However, the proportion change in males was not significant (7% to 6.2%). There was a significant rise in the number of girls being referred for Precocious puberty over 2 decades (3.15% in 2002-03 to 8% in 2022-23).

There was a significant reduction in the proportion of patients referred for Obesity from 14.3% to 7.5% (*P*<0.001). This reduction was significant in both males and females.

Similarly, there was a significant reduction in referrals for disorders of Thyroid gland in both males and females and disorders of Thyroid gland and disorders of Ovaries and Female Reproductive Tract in females (*P*<0.001).

### Comparison of referral reasons with final diagnosis

Out of 2595 patients, 5.3%(n=92) had no pathological findings and investigations were within reference range, thus they were labelled as Normal. This was commonly seen in patients referred for Goitre, Adrenal disorders, Micropenis and Precocious puberty where 28%, 23%, 16.7% and 15% patients had no pathological clinical or biochemical findings, respectively.

The discrepancy between the final diagnosis and referral reason was predominantly observed in patients referred for micropenis, gynaecomastia, and obesity.

Of the 96 patients referred for Micropenis, 42.7%(n=41) had buried penis secondary to Obesity; of the 54 patients referred for Gynaecomastia, 22.2%(n=12) had lipomastia due to Obesity. Of the 257 patients referred for suspected endogenous or secondary Obesity, 220 (85.6%) had exogenous or primary Obesity.

## Discussion

In this retrospective study, we conducted a comprehensive analysis of the reasons for referral to a paediatric endocrine clinic at a tertiary hospital in Western India. We studied the current referral patterns, changes in these patterns over two decades, and the discrepancies between the reasons for referral and the final diagnoses. To the best of our knowledge, this is the first study from India where such an elaborate analysis of all the classes of paediatric endocrine disorders is described. Overall, the most common reason for referral to paediatric endocrine clinic in 2022-23 was Short Stature in both the sexes. This was followed by disorders of Testes and Male Reproductive Tract in males and disorders of Puberty in females. Obesity was the 3rd most common reason for referral in both the sexes. We noticed a substantial increase in the number of referrals to paediatric endocrine clinic from 2002-03 to 2022-23. There was a 1.8-fold increase in total referrals over the span of two decades and the referrals for girls had doubled. The proportion of referrals for Short Stature and disorders of Puberty increased over two decades, particularly in females. A reduction was observed in referral for Obesity and disorders of Thyroid gland in both males and females and in disorders of Ovaries and Female Reproductive Tract. A discrepancy between the final diagnosis and referral reason was predominantly found in patients referred for Micropenis, Gynaecomastia and Obesity.

We found that more than half of the referrals were for Short Stature. Similar trend is reported by a study from Italy where disorders of growth were the most common reason for referral.^[6]^ Among the cases referred for Short Stature at our centre, Idiopathic short stature, Primary growth failure and Secondary growth failure were the most common subcategories in that order. In these, FSS comprised of almost 1/4th of all the cases of Short Stature. Like our findings, a study from Bangladesh has also reported the most common cause of Short Stature as FSS. ^[10]^ Whereas in a study from Northern India and in another from Pakistan, CDGP was reported as the most common cause of Short Stature. ^[11,12]^

There was an increase in the total number of referrals for Short Stature over the two decades. Although the increase was observed in both sexes, the rise was more pronounced in females. This rise could be due to increased awareness among parents and physicians about treatment options, improved screening and growth monitoring tools such as manual or electronic plotting of growth charts, increasing use of automated software tools for growth monitoring, parental expectations and societal pressures, advances in treatment options, early genetic diagnosis of syndromes associated with short stature as well as the modifications in the indication of growth hormone therapy. ^[13,14]^

Among disorders of Puberty, two-thirds of patients were referred for Precocious puberty. Our study identified a significant rise in referrals for precocious puberty in girls between 2002–03 and 2022–23. The earlier onset of puberty, influenced by secular trends, increasing prevalence of obesity, lifestyle changes, and increased exposure to endocrine-disrupting chemicals (EDCs) during and after the COVID-19 pandemic, have been reported in multiple countries, including India. ^[15,16]^ Similarly, a 20-year nationwide study from Denmark documented a significant increase in the annual incidence of central precocious puberty and normal variants of puberty in girls of Danish-origin, with a comparable but less pronounced trend observed in boys. ^[17]^

In children referred for Obesity, Polygenic multifactorial obesity was the most common subcategory. As per a WHO report, the prevalence of obesity among children and adolescents aged 5–19 has risen dramatically from just 2% in 1990 to 8% in 2022. ^[18]^ According to the Indian National and Family Health Survey (NFHS) report, the prevalence of overweight children in India under five years of age has increased from 2.1% (2015–2016) to 3.4% (2019–2021).^[19]^ A meta-analysis of data over 2 decades from India has reported a prevalence of childhood obesity of 8.4%. ^[20]^ However, we found a significant decline in the number of referrals for Obesity over 2 decades but there was a rise in referrals for Micropenis and Gynaecomastia. These were found to be a manifestation of Obesity in half and one third of total referrals for Micropenis and Gynaecomastia respectively. This may be attributed to a lack of awareness and persistent archaic beliefs among parents, coupled with insufficient recognition of the severity of childhood Obesity by primary care paediatricians and physicians. Also continuing medical education may have improved primary health care providers’ knowledge about Obesity, that it is primarily a lifestyle disease and not an endocrine disease in most cases.

Hypothyroidism was the most common reason for referral in disorders of Thyroid Gland. Most of these referrals were already diagnosed or clinically suspected Hypothyroidism in contrary to the other studies where the children were referred for Short Stature and later found to have a Hypothyroidism. ^[12,21]^ This may reflect a lower threshold for ordering a thyroid function test by the primary physician/ paediatrician in children with either Obesity or family history of Thyroid disease. The referrals for disorders of Thyroid Gland have significantly decreased over the past two decades, suggesting that a substantial proportion of these cases are being effectively diagnosed and managed by primary care paediatricians.

Among the disorders of Testes and Male Reproductive Tract, Disorders of penis were the most common referral reason followed by the non-pathological normal variants. However, about 1/5th of patients with Disorders of penis were falsely categorised in the category as Micropenis by referring physicians or paediatricians while the final diagnosis was buried penis, secondary to Obesity.

We noticed a substantial increase in the number of referrals to our paediatric endocrine clinic from 2002-03 to 2022-23, with a 1.5-fold rise in males and more than a two-fold rise in females. A similar trend was reported in an Italian study, where these referrals increased over six years despite a decline in the overall population.^[8]^ The most notable increase was in referrals for disorders of Puberty and Short Stature, with a higher proportion of girls referred for pubertal disorders. No sex bias was identified among children referred for Short Stature. This suggests that parental concerns regarding growth were similar for both sexes, with no significant differences in health-seeking behaviour for short stature observed between boys and girls.

The number of referrals for Ovarian and Female Reproductive Tract Disorders has significantly declined despite a high prevalence of menstrual disorders among adolescent girls in India.^[22,23]^ This shift is likely due to increased referrals to gynaecologists instead of endocrinologists, driven by the greater accessibility to gynaecological speciality, cultural perceptions associating menstrual issues with gynaecology, overlapping expertise, and parental focus on addressing visible menstrual and reproductive health concerns rather than underlying hormonal imbalances.

A discrepancy was observed between the referral reasons and final diagnoses, particularly in patients referred for Micropenis, Gynecomastia, and Obesity. The proportion of Obesity cases misclassified as Micropenis remained consistent over two decades, inappropriately inflating the initial categorization of disorders of the Testes and Male Reproductive Tract. Cases of lipomastia secondary to Obesity are frequently misclassified as Gynecomastia and incorrectly categorized under disorders of Puberty during initial evaluation. Approximately one-fourth of patients referred for Precocious puberty was found to have no pathological findings and were categorized as Normal. Additionally, most Obesity referrals for suspected endocrine causes were ultimately attributed to inappropriate dietary and lifestyle practices. This highlights the need to spread awareness among primary care paediatricians/physicians about presenting complaints of common disorders such as Obesity, where the patient presents with perceived micropenis or acanthosis but has Simple Obesity which needs attention.

This study provides a comprehensive analysis of referral patterns to paediatric endocrine clinics in Western India. It represents the first study to systematically evaluate the evolving referral practices over the past two decades, highlighting disparities between the initial referral indications and the eventual diagnoses. The findings underscore trends in referral practices, offering valuable insights for enhancing the continuing medical education of both primary care providers and specialists. Additionally, the study contributes to optimizing outpatient clinic management in alignment with the current healthcare demands of the population. Furthermore, it identifies critical gaps in the understanding of endocrine disorders, emphasizing the need for targeted educational interventions.

A potential limitation of this study is its retrospective design. The data largely represents the middle or upper middle socioeconomic class population as it is from a hospital catering to this population; the data from a public sector hospital was not taken into consideration. Further, the data are from a single center. However, the main findings on prevalence, change in trend, gender distribution, discrepancy in referral reasons reported in our study are consistent with other studies in the literature.

## Conclusion

We describe recent status and the evolution of patterns of paediatric endocrine referrals over two decades, revealing a significant shift in the number of referrals but not a major shift in the reasons for referrals. Our study also highlights the evolving endocrine disorders with industrial, economic and nutritional transition. A gap exists in recognizing the symptoms and possible cause at the primary care level. These findings emphasize the importance of targeted medical education and awareness among both primary care providers and specialists. While this study highlights these gaps, future research may be directed at the reasons that cause change in the referral pattern over time.

## Data Availability

All data produced in the present study are available upon reasonable request to the authors

